# Impact of the HK-VMT Platform on Early Detection of Cognitive Impairment and Promotion of Healthy Behaviors in Older Adults

**DOI:** 10.1101/2025.05.13.25327489

**Authors:** Fung Ada Wai Tung, Ma Suk Ling

## Abstract

**Background:** The global rise of cognitive impairment and dementia poses significant public health challenges. Existing clinical practice and many social services focused on diagnosis and management after onset. The Hong Kong-Vigilance and Memory Test (HK-VMT) platform combines dementia risk assessment and cognitive test in one accessible tool to enable early detection of dementia in community setting.

**Objective:** This study aimed to evaluate the effectiveness of the HK-VMT platform in assessing dementia risk and a broad spectrum of cognitive impairment in community-dwelling adults. It also assesses the impact of the platform on improving public awareness and encouraging lifestyle changes.

**Methods:** This cross-sectional study assessed 517 adults aged 50 and above recruited through outreach activities between July 2024 and March 2025. Participants underwent a two-stage screening process consisting of dementia risk assessment and cognitive test. The platform collected data on socio-demographic, psychological, medical, and physiological factors for assessing dementia risk using Cognitive Ageing Risk Score (CARS). Cognitive performance was measured by the HK-VMT. User feedback on platform accessibility, adoption, user engagement, public awareness, and attitudes toward healthy lifestyles was obtained through interview.

**Results:** 19.7% of participants were at high risk of dementia. Cognitive impairments were detected in 34.3% of participants through the HK-VMT platform. For user experience, 78% of participants with cognitive impairments were unaware of their condition before screening. Over 95% of participants reported improved understanding of their cognitive health status and over 80% expressed intentions to adopt healthy lifestyle.

**Conclusions:** The HK-VMT platform shows to enhance early detection of cognitive impairments, improve accessibility, increase public awareness and engage the public in brain health management. It represents a scalable solution to support healthy ageing and reduces disparities in early dementia preventive care by bridging community cognitive health services.

## Introduction

Dementia affects 57 million people worldwide in 2021 and is projected to triple by 2050 (1). While the increasing prevalence of dementia is driven by age and genetic factors, increasing evidence suggests that untreated health conditions and poor lifestyle choices are also key contributors to the rise of dementia. The 2024 Lancet Commission report highlighted that nearly half of the incidence could be prevented by addressing 14 modifiable risks, including low education, hearing loss, hypertension, smoking, obesity, depression, physical inactivity, diabetes, excessive alcohol consumption, traumatic brain injury, air pollution, infrequent social contact, untreated vision loss and elevated LDL levels (2). It is important to identify individuals at risk of dementia and to promote a healthy lifestyle early. However, many social centers lack the resources and expertise to conduct a full dementia evaluation, limiting timely interventions and effective allocation of preventive care resource.

To address this gap, the Hong Kong-Vigilance and Memory Test (HK-VMT) platform was launched in mid-2024. The HK-VMT platform is a web application that combines Cognitive Ageing Risk Score (CARS) and HK-VMT to enhance dementia risk assessment and early detection of dementia at the community level. The CARS is a dementia risk assessment tool for estimating the risk of developing dementia based on key risk factors. The HK-VMT, developed in 2021, is a simple and accurate online cognitive screening tool designed to detect early cognitive impairment. The HK-VMT was integrated with CARS in 2024 to become the current HK-VMT platform. Development and validation of both tools have been described elsewhere (3, 4). In brief, the platform incorporates dementia risk assessment to determine individuals who may need further cognitive test. The integration has significantly increased the up-take rate of the HK-VMT from an annual rate of 10% to 82% in 2024, demonstrating the effectiveness of the adoption strategies. It is currently used by 2,047 users from over 20 social centers.

The platform has two separate registration portals for individuals and professionals. Individual users who register through the public portal allows them to link with their health care providers. Once linked, authorized professionals can access and view test results of the linked users, enabling the organization to monitor cognitive status and allocate resources effectively.

The HK-VMT platform is specifically designed to detect subtle cognitive deficits in the preclinical phase to facilitate early intervention and monitoring. The goals of the platform are to provide a free online tool for the public to detect early cognitive changes, raise public awareness of their own dementia risk factors, provide lifestyle advice to reduce dementia risks in high-risk individuals, and support service providers to track cognitive health in their clients.

The current study evaluates the prevalence of dementia risk and the severity distribution of cognitive impairments using the HK-VMT platform. It also evaluates the effectiveness of the platform in detecting early cognitive deficits, raising public awareness of brain health, and promoting healthy behaviors.

## Methods

### Study design and participants

This cross-sectional study was part of an ongoing University knowledge transfer initiative aimed at promoting the HK-VMT platform to support the adoption of healthy lifestyles among high-risk individuals. Between July 2024 and March 2025, a total of 517 participants aged 50 or above were recruited and assessed at community outreach events. The study describes baseline assessment of dementia risk and cognitive deficits. Data collected via the HK-VMT platform included socio-demographic, psychological, cognitive, medical, and physiological factors. Buccal DNA samples were collected for Apolipoprotein E (APOE) genotyping. Cognitive performance was evaluated using HK-VMT. Research assistant administered the Hong Kong version Montreal Cognitive Assessment (HK-MoCA) (5). Additionally, a user feedback survey assessed platform adoption, user engagement, public awareness, and attitudes toward healthy lifestyles. Ethical approval was obtained from the Hong Kong Baptist University Research Ethics Committee (HKBU REC/23-24/0497). Informed consent was obtained from all participants prior to assessment.

#### Description of the HK-VMT Platform

The HK-VMT platform combines CARS and HK-VMT to create a practical two-stage screening process suitable for social settings.

### Stage 1: Dementia risk assessment

CARS estimate the six-year dementia risk based on demographic, health and behavioral risk factors in Chinese older adults, including age, gender, years of education, diabetes, engagement in mind-body exercise, physical inactivity, sleep quality, and level of loneliness (3). A CARS cutoff of −1.3 or above has a sensitivity and specificity of 83.9% and 75.4% for predicting dementia risk. The platform converts these risk factors into survey questions. Four additional dementia risk factors, including hyperlipidemia, hypertension, obesity, and smoking status, are also included to tailor lifestyle advice promoting cognitive and physical activity, weight management, healthy diets, and stress reduction.

### Stage 2: Cognitive assessment

Individuals identified as high risk by CARS would proceed to cognitive testing using the HK-VMT (4). It consists of four subtests assessing attention, psychomotor speed, visuospatial memory, learning and episodic memory. Each subtest generates an individual score that is summed together to form a total score ranging from 0 to 40, with higher scores indicating better cognitive performance. A cut-off of 21/22 differentiates Mild Cognitive Impairment (MCI) from normal cognition with a sensitivity of 86.1% and specificity of 75.3% (4). Raw scores of each subtest are standardized into age- and education-adjusted z-score to calculate the global composite cognitive score.

### Result Dashboard

On completion of the two-stage screening, an immediate comprehensive report is displayed on the result dashboard. The report informs users of their cognitive test score, dementia risk level, likelihood in percentage of developing dementia in six years, attributable modifiable risk factors, and personalized lifestyle recommendations tailored to individual needs. The HK-VMT platform classifies cognitive status into four color-coded categories, with green representing cognitively normal, yellow for cognitive deficits, orange for mild cognitive impairment (MCI), and red for significant impairments suggesting dementia. Users can log in to their personal accounts to access their previous test results and personalized recommendation and to track their cognitive change over time. The platform is accessible free of charge on computers or iPads and is available in Chinese and English.

### Evaluation of the HK-VMT platform

Effectiveness in early detection of dementia was assessed by comparing the distribution identified by the HK-VMT and the HKMoCA. This comparison quantified the platform’s ability to detect previously unrecognized cognitive impairments relative to conventional screening methods. Psychosocial and behavioral outcomes were assessed by measuring public awareness of brain health, identifying barriers to test accessibility, and evaluating attitudes toward healthy lifestyle behaviors.

### Statistical Analysis

Participant characteristics including sociodemographic variables, dementia risk, and their cognitive profile were described using descriptive statistics. Difference between individuals with and without dementia risk on cognitive performance were analyzed using independent t-tests or Chi-square test. Association between modifiable risk factors and cognitive performance was examined using univariate analysis. Data analyses were performed in IBM SPSS Statistics 28.0 for windows. Statistical significance of all analyses was set as p<0.05.

## Results

### Demographic Characteristics

A total of 521 participants were assessed in the study. After excluding 4 participants due to missing data, 517 cases remained for analysis. Of these, 48.2% were enrolled through social centers, while the rest were recruited via public advertisements. The mean age of the sample was 69.3 ± 7.5 years, with 80.1% female participants. Most participants (78.9%) had attained secondary school education. In general, the cohort has a mean CARS score of −3.3 ± 2.5 and a mean HK-VMT score of 27.3 ± 5.1, corresponding to low dementia risk and normal cognitive status. Table 1 provides sample characteristics for 517 participants in the study.

**Table 1.** Demographic Characteristics of the 517 participants.

| **Characteristics** |  | **All (N=517)** |  |
| --- | --- | --- | --- |
|  |  | **Mean (SD)** | **%** |
| Age |  | 69.3 (7.5) | - |
| Females |  | - | 80.1 |
| Education | Years | 10.7 (4.5) |  |
|  | Primary | - | 21.1 |
|  | Secondary | - | 54.7 |
|  | Tertiary | - | 24.2 |
| ApoE4 Genotype | e4 carrier | - | 18.4 |
| CARS |  | -3.3 (2.5) | - |
| HK-VMT |  | 27.3 (5.1) | - |
| HK-MoCA |  | 26.6 (3.6) | - |

### Prevalence of Cognitive Impairment

Cognitive impairments were presented in 34.3% (N=177) participants. Of which, cognitive deficits accounted for 11.4%, MCI accounted for 21.9%, and significant cognitive impairments accounted for the remaining 1%. The prevalence of cognitive impairments increased with age (r=56.4, p<.001). Table 2 shows the overall prevalence for cognitive status stratified by age ranges.

**Table 2.** Prevalence of Cognitive Impairments (in %) classified by HK-VMT.

| **Age** | **CN (N=340)** | **CD (N=59)** | **MCI (N=113)** | **SCI (N=5)** |
| --- | --- | --- | --- | --- |
| 50-59 years | 84.4 (27) | 6.3 (2) | 9.4 (3) | 0 |
| 60-69 years | 76.1 (197) | 9.7 (25) | 14.3 (37) | 0 |
| 70-79 years | 56.6 (99) | 14.3 (25) | 27.4 (48) | 1.7 (3) |
| 80 years | 33.3 (17) | 13.7 (7) | 49 (25) | 3.9 (2) |

CN=Cognitively Normal; CD= Cognitive Deficit; MCI=Mild Cognitive Impairment; SCI Significant Cognitive Impairment

A one-way ANOVA was conducted to compare the cognitive performance of the four cognitive statuses classified by HK-VMT. The results showed that significant differences in the HK-VMT total score (F=235.6.2, p<.001, η2 p=0.579), retrieval (F=97.0, p<.001, η2 p=0..366), retention (F=119.4, p<.001, η2 p=0.415), attention (F=10.1, p<.001, η2 p=0.057), and visuospatial memory (F=64.4, p<.001, η2 p=.277), based on the cognitive status stratified by CARS.

HK-MoCA identified only 10.1% of participants as having MCI. In contrast, the HK-VMT platform identified 34.3% of the participants with cognitive impairment. Of those identified as having MCI, 15.9% (N=82) were not detected by the HK-MoCA.

### Prevalence of Dementia Risk Factors and Association with Cognition

A majority of the participants (98%) had at least one modifiable risk factor. Of which, 19.7% were classified by CARS as having a high risk of dementia. The most common risk factors were poor sleep (67.7%), loneliness (65.2%), and hypertension (50.1%). Other risk factors included hyperlipidemia (23%), diabetes (13.5%), overweight or obesity (10.1%), and smoking (0.4%). The sample was relatively active, with only 4.8% lacking physical activities, but 48.9% did not engage in mind-body exercises.

Each modifiable risk factor was further examined individually with cognitive performance. Table 3 displayed the difference in HK-VMT score between those with and without a particular risk factor. The HK-VMT score was lower in individuals with diabetes, hypertension, no engagement in mindbody exercise, poor sleep, and loneliness than those without such risk factors. We further analyzed whether ApoE e4 carrier would modify the association between modifiable risk factors and cognitive performance. Univariate analysis demonstrated interaction between ApoE4 and hypertension is significantly associated with cognitive performance (F(5,449)=3.09, p=.047).

**Table 3.** Comparison of HK-VMT score in individuals with and without dementia risk factors.

| Risk Factors | Presence |  | Absence |  | T-test |
| --- | --- | --- | --- | --- | --- |
|  | Mean | SD | Mean | SD | *p*-value |
| Diabetes | 25.2 | 5.4 | 27.6 | 5.0 | *** |
| Hypertension | 27.3 | 4.9 | 28.6 | 4.5 | ** |
| Hyperlipidemia | 28.3 | 4.9 | 28.2 | 4.9 | ns |
| No Mindbody Exercise | 26.6 | 5.0 | 27.9 | 5.2 | ** |
| Physical inactivity | 27.8 | 5.4 | 27.2 | 5.1 | ns |
| Overweight/Obesity | 27.7 | 6.2 | 27.3 | 5.0 | ns |
| Poor sleep | 26.9 | 5.2 | 28.0 | 4.9 | ** |
| Loneliness | 27.0 | 5.2 | 27.9 | 5.0 | * |

\*\*\* $<.001$ ; \*\* $<.01$ , \* $<.05$ , ns=not significant, ApoE4= Apolipoprotein e4. S.D.= Standard deviation.

### Impacts on Awareness and Motivation for Behavioral Change

78% (N=138) of participants with cognitive impairments were unaware of their condition before screening. Furthermore, the platform played a significant role in raising public awareness and encouraging positive behavioral changes. Following the screening, over 95% of participants reported improved understanding of their cognitive health status with a knowledge rating of 4 out of 5, indicating satisfactory level of awareness. A substantial proportion of participants with cognitive impairments expressed intentions to adopt healthier lifestyle, with 87% planned to increase their physical activity, 77% intended to improve their diet, and 70% aimed to pay closer attention to brain health information; 58% considered undergoing regular checkup on their cardiovascular health; and 54% planned to engage more volunteering or social activities.

Qualitative feedback highlighted increased awareness, motivation for lifestyle changes, and a desire for regular cognitive health monitoring. Participant quotes reflected appreciation for the platform’s accessibility and its role in raising public awareness and motivating behavior change:

- “V*ery helpful, especially for community older people who generally have limited access to such information*”. (BHD480)
- “*The screening helped me to gain a deeper understanding of the knowledge about memory and alertness. I can also refer to the report to improve my poor habits*.” (BHD403)
- “*Thank you so much for conducting this assessment on me! It has given me a much deeper understanding of cognitive impairment, which will be incredibly helpful for my future lifestyle and healthy living habits*.”. (BHD429)
- “*It’s really great! It allows for an initial assessment of my cognitive abilities and health, which helps me to make improvement. I hope it can be done regularly, like once a year, for screening purposes*.”. (BHD414)

## Discussion

This cohort study addresses the information gap regarding early cognitive deficits among older adults in the community. The challenge lies in distinguishing subtle cognitive changes from normal ageing. Currently, preclinical cases with subtle cognitive deficits but do not meet the criteria for MCI can only be verified through sophisticated neurological examination looking for biomarkers related to brain pathology. To our knowledge, HK-VMT platform is the first assessment tool that enables early detection preceding MCI in social setting. Over the past decade, many cognitive screening tools have primarily focused on identifying MCI without providing personalized recommendations (6-12). Recent online risk assessment tools, such as CogDrisk or LinusHealth, provide risk evaluations but do not integrate cognitive test results into their dementia risk analyses. In contrast, our platform serves as a comprehensive dementia risk management system that tailors assessments and recommendations to each individual’s cognitive profile.

Our findings show that 34.3% of adults aged 50 or above experience varying levels of cognitive impairment in Hong Kong. Of which, 21.9% were classified as having MCI. This is comparable to previous epidemiological estimates of 21.2%, demonstrating the platform’s validity in detecting MCI in local population (13). The implementation of the platform also allowed us to increase detection rate of MCI by 15.9% when compared to conventional screening. Notably, over 80% of those unidentified MCI cases attained secondary school education or higher, suggesting that the platform is particularly suitable for highly educated individuals.

More importantly, we are the first to report the prevalence of cognitive deficits detected in social setting. The HK-VMT platform uses a two-stage screening process that stratifies cognitive profile by dementia risk. This approach reveals that over 10% of participants exhibit cognitive deficits with multiple modifiable risk factors. Consistent with previous work, individuals with cognitive deficits exhibit a significant drop in attention and learning (3). The findings can inform population-based preventive strategies aimed at enhancing support for individuals with hypertension, diabetes, poor sleep, loneliness, as well as promoting mind-body exercise to lower the rise of cognitive impairment. Furthermore, our result indicated that genetic predisposition modifies the relationship between hypertension and cognitive outcomes, suggesting individuals with a family history of dementia should engage in early blood pressure monitoring and adopt healthy lifestyle practice to support brain health. This new information provided by the platform empowers high-risk individuals to take proactive step for dementia risk reduction.

Following on these findings, our study also revealed that a significant proportion (nearly 80%) of individuals were unaware of their condition before screening. This widespread unawareness indeed reflects a lack of resources for early screening in the community. In Hong Kong, clinical care primarily focuses on diagnosis and treatment after dementia onset, while social centers provide welfare services for older adults with cognitive or functional decline. Since 2019, District Health Centres (DHCs) and DHC Expresses (DHCEs) have provided subsidized primary healthcare for individuals with various health conditions. However, these community services lack expertise and device to provide comprehensive dementia risk assessments for early detection. As a result, many older people at risk of dementia miss out on early preventive care. Indeed, our findings showed that older people who are unaware of their cognitive are highly motivated to make changes in adopting a more active and healthier lifestyle. These findings suggest that the platform does not only raise awareness but also motivates individuals to engage in healthier behaviors. This suggests that the integration of the HK-VMT platform into routine community screening offers an efficient method for early identification of cognitive impairment in older adults.

The present study had several limitations. First, this is a cross-sectional study estimating the prevalence of dementia risk and cognitive impairments using the HK-VMT platform, and thus it cannot assess the progression rate of dementia risk and the incidence of dementia. Second, most of our participants were females and with higher education, the result may not be applicable to males and those with less education. Third, the causal relationship between platform usage and cognitive change could not be determined due to cross-sectional design. Yet, this digital screening initiatives provides solid data for future tracking on community brain health. Overall, the study demonstrates the value of the platform in empowering users through knowledge and encouraging lifestyle modifications that may improve cognitive outcomes in the long run.

## Conclusion

Early identification is critical for effective dementia prevention. The HK-VMT platform demonstrates significant potential to enhance early detection, accessibility, and public engagement in cognitive health among older adults. Its high adoption rate and the ability to detect cognitive deficits prior to MCI highlights its suitability for large-scale community screening. The initiative exemplifies a scalable approach to reduce disparities in preventive cognitive care.

## Data Availability

All data produced in the present work are contained in the manuscript.

## Acknowledgments

The authors thank the research assistants, Miss Able Kwok Cheuk Lam, Mr. Dicky Tik Man Leung, Mr. Michael Cheuk Yin Chu, and Mr. Kin Chung Wong for their assistance in data collection. Also, the authors expressed gratitude to the Hong Kong Red Cross, the New Life Church of Christ Sun Tin Wai Neighbourhood Elderly Centre, Hong Kong Christian Mutual Improvement Society Ko Chiu Road Centre of Christ Love for the Aged, and all participating organizations and older adults for tremendous support and participation throughout the study.

## Funding

This study was funded by the Research Network of Healthy Ageing under the Faculty of Arts and Social Science, Hong Kong Baptist University. (Ref: RNHA202302)

## Conflicts of Interest

Nil.

## Notes

### Competing Interest Statement

The authors have declared no competing interest.

### Author Declarations

Ethical approval was obtained from the Hong Kong Baptist University Research Ethics Committee (HKBU REC/23-24/0497).

